# Towards Interpretable Risk: Multidimensional Context for ICU Mortality Predictions

**DOI:** 10.64898/2026.09.03.26362203

**Authors:** Shraddha Gupta, Akanta Das, Mrinmoy Sarkar Anto, Zamiul Alam, Arnob Datta, Tanmay Gupta, Tanmoy Sarkar Pias, Humayera Islam

## Abstract

ICU mortality models can achieve strong discrimination, yet a risk score alone provides limited context for patient-level interpretation. We developed a multidimensional prediction-context framework that complements a calibrated mortality estimate with model behavior, data availability, recent physiology, and model attribution. Using 3,236 held-out ICU episodes from the MIMIC-III in-hospital mortality benchmark, we compared LSTM, GRU-D, XGBoost, and a weighted ensemble. The ensemble achieved an AUROC of 0.871 and AUPRC of 0.536; validation-based logistic recalibration improved the Brier score from 0.134 to 0.075. Incorrect predictions showed greater component-model disagreement and smaller decision-boundary margins, although model agreement and large margins did not guarantee correctness. Observation coverage and recent physiological trends also varied substantially across patients, highlighting differences in the information surrounding otherwise similar risk estimates. SHAP analysis attributed 89.9% of total absolute XGBoost attribution to physiological-value features and 10.1% to observation-process features. These dimensions were integrated into patient-level profiles to provide a more complete view of how predictions were formed and the clinical and data context surrounding them, extending interpretation beyond risk scores and feature rankings alone. Abbreviations: ICU (Intensive care unit)

## 1. Introduction

In-hospital mortality prediction is a longstanding problem in critical care informatics. Conventional severity scores such as APACHE, SAPS, and SOFA remain widely used, but they summarize illness severity from relatively limited clinical snapshots and do not fully represent the evolving physiological trajectories captured in electronic health records (EHRs)^1–3^. Machine-learning (ML) and deep-learning (DL) models can leverage longitudinal EHR data and achieve strong discrimination^4–5^, but discrimination alone is insufficient for patient-level decision support. A predicted probability must also be appropriately calibrated and interpreted in relation to the patient’s current clinical state^6–7^. Importantly, two patients with similar predicted risks may differ substantially in model agreement, recent data availability, physiological trajectory, and the information driving the prediction.

Explainable artificial intelligence has therefore become increasingly common in critical-care prediction. SHapley Additive exPlanations (SHAP) and related attribution methods can identify which variables contributed to a model output^8^, and several ICU mortality studies now provide global and patient-level explanations^9–10^. However, explaining what influenced a prediction is not the same as providing the context needed to interpret that prediction. A ranked feature list does not indicate whether component models agree, whether the prediction lies near the operating threshold, whether important measurements were recently available, or whether the patient’s physiology is stable or changing. Although calibration, model disagreement, missingness, temporal dynamics, and feature attribution have each been studied, these elements are typically addressed separately rather than presented as context surrounding the same patient-level prediction^10–12^.

This distinction is particularly important in ICU EHR data because the record reflects both patient physiology and the clinical observation process^13^. Measurement frequency and missingness are often informative rather than random^13–15^, meaning that model attribution may reflect not only physiological values but also how frequently, recently, or whether a variable was measured. Consequently, feature importance alone may provide an incomplete representation of the evidence surrounding a prediction. More broadly, model agreement or distance from a decision threshold should not be assumed to represent reliability, just as post-hoc attribution should not be interpreted as a guarantee of prediction correctness^16^.

To address this gap, we propose a multidimensional prediction-context framework centered on a calibrated patient-level mortality probability and organized around four complementary dimensions: model context, describing component-model behavior and decision-boundary position; data context, describing recent measurement availability and coverage; recent physiological context, describing observed values and trajectories; and explanation context, describing what information contributed to the model output and whether that contribution arose from physiological values or the observation process. Our contribution is a structured approach that extends prediction explanation to prediction contextualization by showing not only what the model predicted, but also how the prediction was formed, what information was available, what the patient’s recent physiology looked like, and what drove the model output. We evaluate whether these dimensions provide distinct and complementary information around ICU mortality predictions and demonstrate how they can be integrated into patient-level profiles as a foundation for subsequent clinician-centered evaluation.

## 2. Methods

### 2.1 Study design and data source

We conducted a retrospective cohort study using the MIMIC-III^17^ in-hospital mortality benchmark^4^, comprising adult ICU episodes from Beth Israel Deaconess Medical Center with a standardized 48-hour observation window across 17 benchmark variables. The study evaluated a multidimensional framework for contextualizing patient-level mortality predictions. We retained the official benchmark partition of 14,681 training, 3,222 validation, and 3,236 held-out test episodes. All model development, ensemble-weight selection, and probability calibration were performed without accessing test-set labels. Observed in-hospital mortality in the train set was 13.5% and in the test set, 11.6%. Model training and ensemble selection were performed using the training and validation sets, and the held-out test set was reserved for final evaluation. Figure 1 summarizes the overall framework.

**Figure 1.**
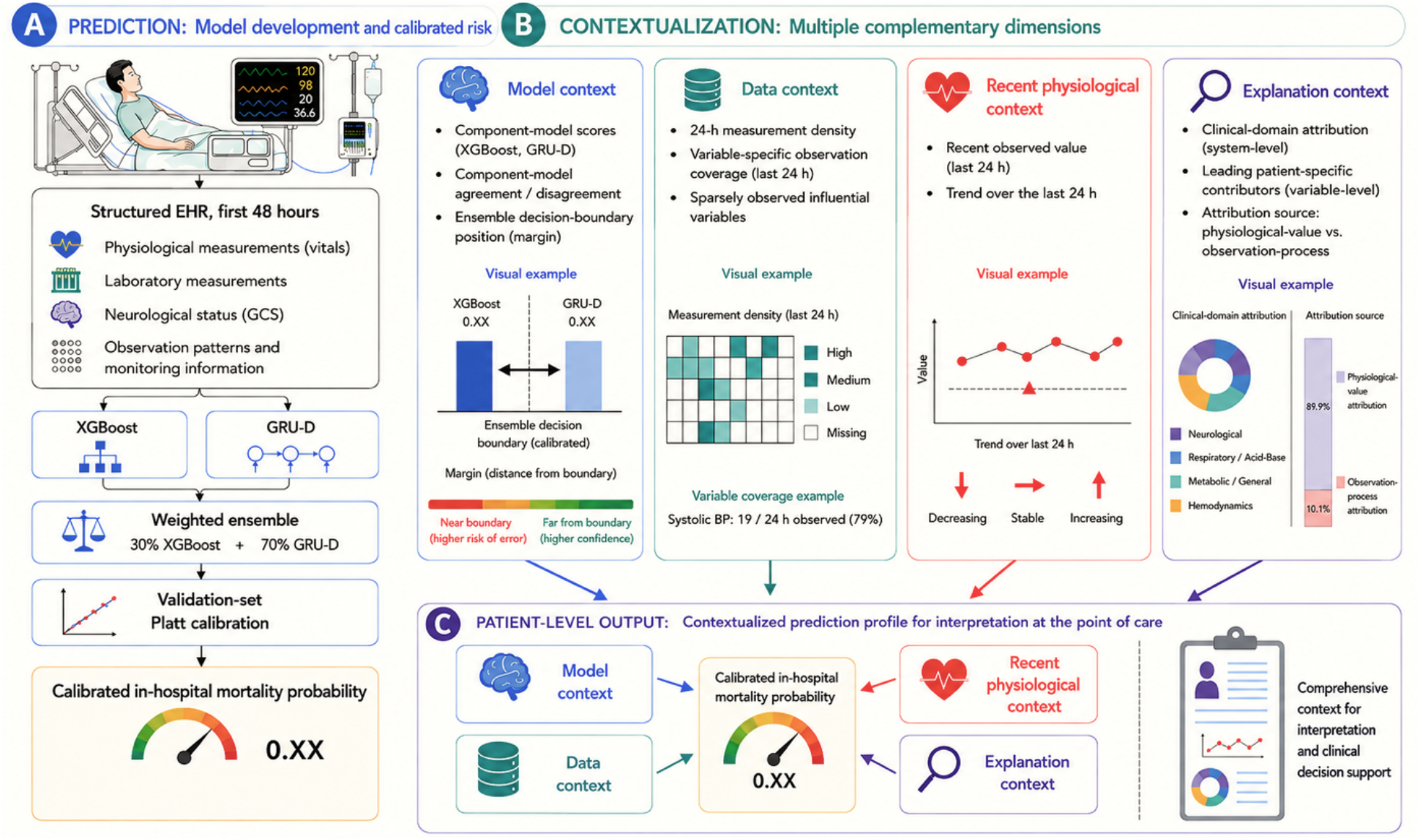
Multidimensional framework for contextualizing ICU mortality predictions. The framework combines a calibrated ensemble mortality probability with complementary model, data, recent physiological, and explanation context to support patient-level interpretation without modifying the original prediction.

### 2.2 Input representation and model development

Each ICU episode was represented as a 48-by-17 hourly matrix of normalized clinical measurements. The recurrent models operated directly on this longitudinal representation, whereas XGBoost used a fixed-length engineered feature representation derived from the same 48-hour data. Normalization parameters were estimated from observed training values only. For the LSTM, normalized measurements were concatenated with binary observation masks at each time step. GRU-D used the 17-variable time series together with observation masks and time since last measurement, with learned feature-wise decay toward the empirical mean and hidden-state decay to model irregularly observed data.^14^. For XGBoost, seven summary statistics (mean, standard deviation, minimum, maximum, skewness, count, and missingness indicator) were computed across seven temporal windows for each of the 17 variables, yielding 833 candidate features. Seven height features were unavailable across these temporal windows, resulting in 826 features. Remaining missing values were imputed with training-set column means.

We evaluated LSTM, GRU-D, and XGBoost^18^. The LSTM used 64 hidden units and dropout 0.3. GRU-D used 256 hidden units, dropout 0.2, Adam optimization (learning rate 5 × 10^−4^, weight decay 10^−5^), learning-rate reduction on plateau, and early stopping. XGBoost used 1,000 estimators, maximum depth 4, learning rate 0.02, row and column subsampling of 0.8, class weighting, and validation-based early stopping.

The GRU-D/XGBoost ensemble weights (*w*′*s*) were selected by validation-set grid search over {0.00, 0.05, …, 1.00}, optimizing AUPRC, yielding:

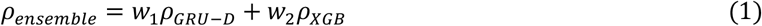

For the clinical-domain analysis, models were retrained using variables belonging to four clinical systems: Hemodynamics, Respiratory/Acid-Base, Neurological, and Metabolic/General over the full 48-hr window. For the temporal-window analysis, all available variables were retained while the observation history was restricted to the final 12 hours, final 24 hours, or complete 48-hour benchmark window. Models were retrained separately for each temporal window using condition-specific preprocessing. Performance was compared using AUROC, AUPRC, accuracy, balanced accuracy, positive-class precision, recall, and F1 score (Table 1).

**Table 1.** Predictive performance across clinical-domain subsets and temporal input windows. Metrics include area under the receiver operating characteristic curve (AUROC), area under the precision-recall curve (AUPRC), accuracy (Acc), balanced accuracy (BAcc), positive-class precision (Prec1), recall (Rec1), and F1 score (F11) using a threshold of 0.50. Bold values indicate the strongest ensemble performance within the corresponding experimental setting.

| Setting | Model | AUROC | AUPRC | Acc | BAcc | Prec<sub>1</sub> | Rec<sub>1</sub> | F1<sub>1</sub> |
| --- | --- | --- | --- | --- | --- | --- | --- | --- |
| **Clinical Domain Subsets (Full 48h Window)** |  |  |  |  |  |  |  |  |
| Hemodynamics | LSTM | 0.664 | 0.276 | 0.543 | 0.602 | 0.158 | 0.679 | 0.256 |
|  | GRU-D | 0.697 | 0.289 | 0.599 | 0.631 | 0.176 | 0.674 | 0.280 |
|  | XGBoost | 0.698 | 0.274 | 0.746 | 0.633 | 0.224 | 0.487 | 0.307 |
|  | Ensemble | 0.709 | 0.303 | 0.659 | 0.649 | 0.197 | 0.636 | 0.301 |
| Resp./Acid-Base | LSTM | 0.729 | 0.317 | 0.537 | 0.651 | 0.174 | 0.800 | 0.285 |
|  | GRU-D | 0.733 | 0.318 | 0.634 | 0.670 | 0.199 | 0.717 | 0.312 |
|  | XGBoost | 0.778 | 0.349 | 0.742 | 0.692 | 0.252 | 0.626 | 0.359 |
|  | Ensemble | 0.782 | 0.360 | 0.729 | 0.693 | 0.245 | 0.647 | 0.355 |
| **Neurological** | LSTM | 0.789 | 0.351 | 0.735 | 0.727 | 0.263 | 0.717 | 0.384 |
|  | GRU-D | 0.795 | 0.359 | 0.730 | 0.727 | 0.260 | 0.722 | 0.382 |
|  | XGBoost | 0.809 | 0.359 | 0.755 | 0.736 | 0.280 | 0.711 | 0.402 |
|  | **Ensemble** | **0.813** | **0.373** | **0.755** | **0.735** | **0.279** | **0.709** | **0.400** |
| Metabolic/General | LSTM | 0.676 | 0.222 | 0.673 | 0.630 | 0.193 | 0.575 | 0.289 |
|  | GRU-D | 0.717 | 0.253 | 0.698 | 0.656 | 0.213 | 0.602 | 0.315 |
|  | XGBoost | 0.716 | 0.257 | 0.730 | 0.646 | 0.223 | 0.537 | 0.315 |
|  | Ensemble | 0.731 | 0.273 | 0.727 | 0.663 | 0.230 | 0.580 | 0.330 |
| **Temporal Window Analysis** |  |  |  |  |  |  |  |  |
| Last 12h | LSTM | 0.839 | 0.470 | 0.761 | 0.768 | 0.296 | 0.778 | 0.429 |
|  | GRU-D | 0.845 | 0.483 | 0.749 | 0.771 | 0.288 | 0.800 | 0.424 |
|  | XGBoost | 0.853 | 0.487 | 0.791 | 0.762 | 0.321 | 0.725 | 0.445 |
|  | Ensemble | 0.850 | 0.496 | 0.757 | 0.775 | 0.296 | 0.797 | 0.432 |
| Last 24h | LSTM | 0.841 | 0.475 | 0.791 | 0.757 | 0.319 | 0.714 | 0.441 |
|  | GRU-D | 0.862 | 0.514 | 0.783 | 0.778 | 0.319 | 0.770 | 0.451 |
|  | XGBoost | 0.852 | 0.487 | 0.794 | 0.754 | 0.321 | 0.701 | 0.440 |
|  | Ensemble | 0.864 | 0.523 | 0.791 | 0.780 | 0.327 | 0.765 | 0.458 |
| Full 48h (ref.) ★ | LSTM | 0.853 | 0.500 | 0.788 | 0.770 | 0.321 | 0.746 | 0.449 |
|  | GRU-D | 0.864 | 0.521 | 0.787 | 0.780 | 0.323 | 0.770 | 0.455 |
|  | XGBoost | 0.865 | 0.524 | 0.806 | 0.769 | 0.340 | 0.722 | 0.462 |
|  | Ensemble | **0.871** | **0.536** | **0.793** | **0.782** | **0.330** | **0.767** | **0.462** |

### 2.3 Probability calibration

For the AUROC and AUPRC of the ensemble model, 95% confidence intervals were estimated using nonparametric 1000 bootstrap resamplings of the test cohort. Probability accuracy was assessed using the Brier score^21^. Because the raw weighted-ensemble scores overestimated absolute mortality risk, Platt scaling^19–20^ was fitted using the validation set only. The fitted transformation was

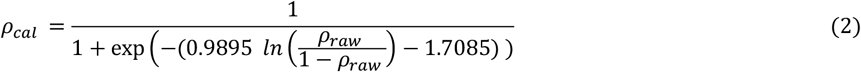

The fitted calibration model was then applied without refitting to the held-out test set. Calibration was evaluated using the Brier score, calibration intercept, calibration slope, and mean predicted probability^6,21^ relative to observed mortality (Table 2). The original ensemble operating threshold of 0.50 was transformed through the validation-fitted calibration model, yielding an equivalent calibrated probability threshold of 0.1534 (used for ensemble classification and decision boundary analyses).

**Table 2.** Held-out discrimination and probability calibration of the raw and calibrated ensemble (n = 3,236). Bootstrap 95% CIs are shown for the raw and calibrated ensemble. Operating threshold: raw 0.500, calibrated 0.1534.

| Model output | AUROC (95% CI) | AUPRC (95% CI) | Brier score | Mean predicted probability | Calibration intercept | Calibration slope |
| --- | --- | --- | --- | --- | --- | --- |
| Raw ensemble | 0.871 (0.854-0.888) | 0.536 (0.485-0.588) | 0.134 | 0.307 | -1.829 | 0.978 |
| Calibrated ensemble | **0.871 (0.854-0.888)** | **0.536 (0.485-0.588)** | **0.075** | **0.125** | **-0.140** | **0.989** |

### 2.4 Model-context analysis

#### Component-model disagreement

For each episode, disagreement between the component probabilities of the ensemble model for ICU-stay *i* was defined as

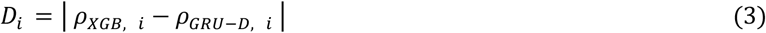

For descriptive presentation, disagreement was grouped as low (*D* < 0.08), moderate (0.08 ≤ *D* < 0.20), or high (*D* ≥ 0.20). The proportion of the held-out cohort and retrospective ensemble error rate were calculated within each category. Disagreement distributions were also compared between correct and incorrect ensemble predictions (Figure 2 (1A)).

**Figure 2.**
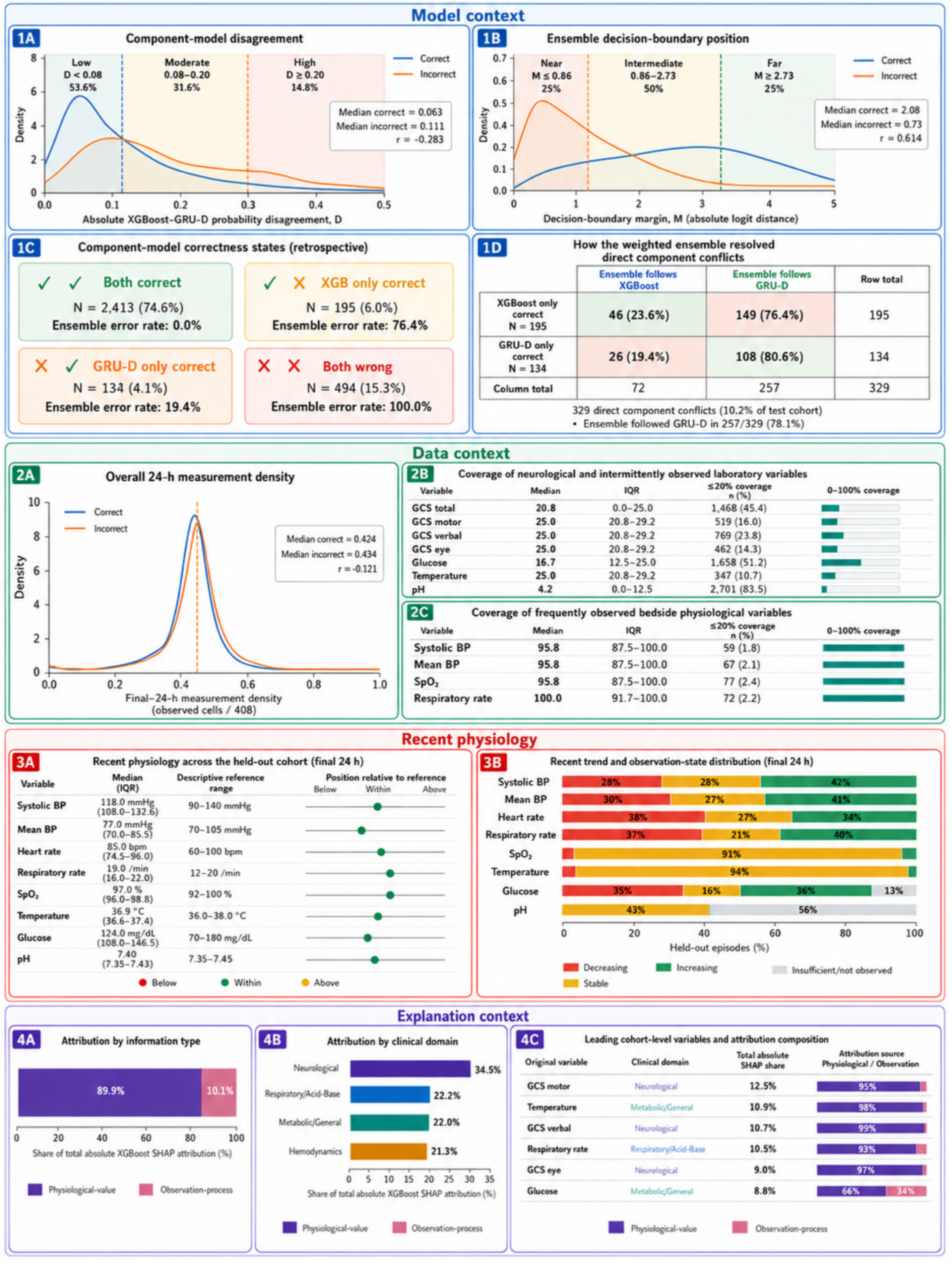
Multidimensional contextualization of calibrated ICU mortality predictions in the held-out test cohort. Model context summarizes component disagreement, decision-boundary position, component correctness states, and conflict resolution; data context describes recent measurement density and variable-specific coverage; recent physiological context summarizes observed values and trends; and explanation context characterizes XGBoost attribution by information type, clinical domain, and leading variables. (*Rank-biserial correlation: r* < 0 *indicates larger values among correct predictions & r* > 0 *indicates larger values among incorrect predictions.)*

#### Decision-boundary position

The signed distance between the calibrated ensemble probability and calibrated operating threshold for ICU-stay *i* was calculated on the logit scale:

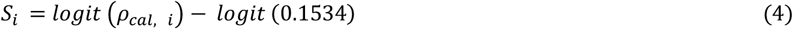

The absolute decision-boundary margin for ICU-stay *i* was

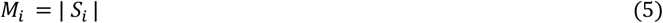

For descriptive visualization, the held-out cohort distribution was divided using the first and third quartiles of the margin. Predictions in the lowest quartile were labeled near the operating boundary, the middle 50% intermediate, and the highest quartile far from the operating boundary (Figure 2 (1B)).

#### Component-model states and direct conflicts

Each test episode was retrospectively assigned to one of four component-model correctness states: both models correct, XGBoost only correct, GRU-D only correct, or both models wrong (Figure 2 (1C)). We additionally identified episodes in which XGBoost and GRU-D made opposite binary decisions. Within this direct-conflict subset, we determined whether the weighted ensemble followed the XGBoost or GRU-D decision and whether that decision was correct (Figure 2 (1D)).

### 2.5 Data-context analysis

Data context was evaluated using the final 24 hours of the 48-hour model input period. Observation status was determined from the masks, while physiological values were obtained from the original test time-series data. Overall, 24-hour measurement density was defined as:

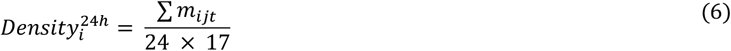

where *i* denotes the ICU episode, *j* the clinical variable, *t* the hourly interval, and *m_ijt_* = 1 when *j* was observed for episode *i* at hour *t*, 0 otherwise. Density distributions were compared between correct and incorrect ensemble predictions (Figure 2 (2A)). Variable-specific coverage was calculated as the percentage of the final 24-hour intervals in which each variable was observed. A variable was described as sparsely observed when coverage was 20% or less. For presentation, variable-level coverage was separated into neurological and intermittently observed laboratory measures (GCS total, GCS motor, GCS verbal, GCS eye, glucose, temperature, and pH) and frequently observed bedside physiological measures (systolic blood pressure, mean blood pressure, oxygen saturation, and respiratory rate). For each variable, median coverage, interquartile range, and the number and percentage of episodes with sparse observation were summarized (Figure 2, 2B–2C).

### 2.6 Recent physiological context

Recent physiology was summarized using the median observed value during the final 24 hours for systolic blood pressure, mean blood pressure, heart rate, respiratory rate, oxygen saturation, temperature, glucose, and pH. Cohort-level medians and interquartile ranges were displayed relative to predefined descriptive reference intervals: systolic blood pressure 90–140 mmHg, mean blood pressure 70–105 mmHg, heart rate 60–100 beats/min, respiratory rate 12– 20/min, oxygen saturation 92%–100%, temperature 36.0–38.0°C, glucose 70–180 mg/dL, and pH 7.35–7.45. (Figure 2, (3A)). Recent temporal trends were estimated from observed-only values using a linear trajectory when at least two measurements were available. Relative fitted change: 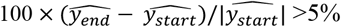 was classified as increasing, <−5% as decreasing, and otherwise as stable; fewer than 2 as insufficient (Figure 2, (3B)).

### 2.7 Explanation context

SHAP values for the XGBoost component were computed using TreeExplainer. Each feature was mapped to one of four clinical domains (Hemodynamics, Respiratory/Acid-Base, Neurological, and Metabolic/General), and an information type. Features summarizing measured clinical values were classified as physiological-value features, whereas count and missingness-related features were classified as observation-process features. Absolute SHAP values were aggregated within each grouping to quantify cohort-level attribution (Figure 2, 4A–4C).

### 2.8 Patient-level prediction profiles

Four held-out episodes were selected retrospectively for illustration: one true positive, one true negative, one false positive, and one false negative. The cases were selected to demonstrate how the different contextual dimensions could be presented around an individual calibrated mortality probability. Each patient profile combined the calibrated mortality probability with component-model scores and agreement, decision-boundary position, clinical-domain attribution, and the four leading patient-specific XGBoost contributors. For each contributor, the display summarized attribution magnitude and direction, recent physiology and trend, observation frequency, and physiological-value versus observation-process contribution.

### 2.9 Statistical analysis

Continuous variables were summarized using medians and interquartile ranges, and categorical variables using counts and percentages. Differences in disagreement, decision-boundary margin, and measurement density between correct and incorrect predictions were assessed using two-sided Mann-Whitney U tests, with rank-biserial correlation^22^ reported as the effect size. Statistical significance was defined as two-sided *p* < 0.05. Analyses were performed in Python using SciPy, scikit-learn, NumPy, pandas, SHAP, PyTorch, Matplotlib, and XGBoost.

## 3. Results

### 3.1 Predictive performance across models, clinical domains, and temporal window experiments

The weighted ensemble, assigned 70% weight to GRU-D and 30% to XGBoost, achieved the highest discrimination, with an AUROC of 0.871 and an AUPRC of 0.536, compared with 0.865/0.524 for XGBoost, 0.864/0.521 for GRU-D, and 0.853/0.500 for LSTM (Table 1). The ensemble also maintained the highest balanced accuracy (0.782) and matched XGBoost for the highest F1 score (0.462). We selected the ensemble model for the subsequent contextual analyses.

The clinical-domain experiments showed that predictive information was distributed across all four physiological domains, although performance differed substantially by domain (Table 1). The neurological subset produced the highest isolated-domain discrimination, reaching an AUROC of 0.813 and AUPRC of 0.373. Respiratory/Acid-Base information provided the next strongest discrimination (ensemble AUROC 0.782, AUPRC 0.360), whereas the Hemodynamics and Metabolic/General subsets produced lower performance.

Temporal-window experiments similarly showed that substantial predictive information was retained in the most recent portion of the ICU stay. Using only the final 24 hours, the ensemble achieved an AUROC of 0.864 and AUPRC of 0.523, close to the full 48-hour performance of 0.871 and 0.536. With only the final 12 hours, ensemble performance decreased to an AUROC of 0.850 and AUPRC of 0.496. XGBoost produced a slightly higher AUROC than the ensemble in the 12-hour experiment (0.853 vs 0.850), whereas the ensemble retained the highest AUPRC.

### 3.2 Held-out discrimination and probability calibration

On the held-out test cohort of 3,236 ICU episodes, the raw ensemble probabilities substantially exceeded the observed mortality prevalence: the mean raw predicted probability was 0.307 compared with an observed mortality rate of 0.116. The raw Brier score was 0.134, with a calibration intercept of −1.829 and slope of 0.978. Validation-set Platt calibration substantially improved probability calibration without changing rank-based discrimination. After calibration, the mean predicted probability decreased to 0.125, and the Brier score improved to 0.075. The calibration intercept improved to −0.140 and the slope to 0.989, while AUROC and AUPRC remained 0.871 and 0.536, respectively (Table 2). The original raw operating threshold of 0.50 corresponded to a calibrated mortality probability of 0.153.

### 3.3 Model context: disagreement, decision-boundary position, and ensemble behavior

Component-model disagreement was lower among correct than incorrect ensemble predictions, with median disagreement of 0.063 versus 0.111, respectively (rank-biserial *r* = −0.283, *p* < 0.001; Figure 2 (1A)). Error rates also increased across the descriptive disagreement categories, from 14.3% in the low-disagreement group to 24.1% with moderate disagreement and 36.5% with high disagreement. Correct predictions were farther from the calibrated operating boundary than incorrect predictions, with median absolute logit margins of 2.080 and 0.731, respectively (*r* = 0.614, *p* < 0.001; Figure 2 (1B)). In the held-out cohort, the lowest quartile of margins was classified descriptively as near the operating boundary (*M* ≤ 0.86), the middle 50% as intermediate, and the upper quartile as far from the boundary (*M* ≥ 2.73). The individual predictions from the XGBoost and GRU-D components in the ensemble model were both correct for 2,413 episodes (74.6%), whereas both were wrong for 494 (15.3%) (Figure 2 (1C)). The components made opposite binary decisions in only 329 episodes, representing 10.2% of the test cohort. Among these conflicts, XGBoost alone was correct in 195 episodes, and GRU-D alone was correct in 134. Consistent with the assigned 70% weight to GRU-D, the ensemble followed the GRU-D decision in 257 of the 329 conflicts (78.1%) (Figure 2 (1D)). Consequently, the ensemble was correct in 80.6% of episodes in which GRU-D alone was correct but in only 23.6% of episodes in which XGBoost alone was correct.

### 3.4 Data context: recent measurement availability

Overall final-24-hour measurement density differed only modestly between correct and incorrect predictions. Median density was 0.424 among correct predictions and 0.434 among incorrect predictions (*r* = −0.121, *p* < 0.001; Figure 2 (2A)), indicating only a small association between overall measurement density and prediction correctness. Variable-specific coverage, however, showed marked differences in how frequently different types of clinical information were available (Figure 2, (2B-2C)). Among neurological and intermittently measured laboratory variables, median final-24-hour coverage was 25.0% for the GCS motor, verbal, and eye components, 20.8% for total GCS, 16.7% for glucose, and only 4.2% for pH. Sparse observation, defined as coverage of 20% or less, occurred in 83.5% of episodes for pH, 51.2% for glucose, and 45.4% for total GCS. In contrast, bedside physiological variables were observed much more consistently. Median coverage was 95.8% for systolic blood pressure, mean blood pressure, and oxygen saturation and 100% for respiratory rate, with sparse observation occurring in only 1.8%-2.4% of episodes.

### 3.5 Recent physiological context

Recent physiological summaries were generated from observed values during the final 24 hours of the input window (Figure 2, Recent physiology). Across the held-out cohort, the median cohort-level values for systolic blood pressure, mean blood pressure, heart rate, respiratory rate, oxygen saturation, temperature, glucose, and pH fell within their descriptive reference ranges (Figure 2 (3A)). For example, the cohort median systolic blood pressure was 118 mmHg (IQR, 108-133), respiratory rate was 19/min (IQR, 16-22), glucose was 124 mg/dL (IQR, 108-147), and pH was 7.40 (IQR, 7.35-7.44). Moreover, recent trajectories showed considerably more heterogeneity than the cohort medians alone (Figure 2 (3B)). Systolic and mean blood pressure were increasing in approximately 42% of episodes and decreasing in 28%-30%, while respiratory rate increased in 40% and decreased in 37%. By contrast, oxygen saturation and temperature were predominantly stable, at 91% and 94%, respectively. Availability also affected whether a recent trajectory could be estimated: 55.7% of pH series and 12.8% of glucose series had insufficient recent observations or no observation for trend estimation.

### 3.6 Explanation context

XGBoost attribution was predominantly driven by physiological-value features, which accounted for 89.9% of total absolute SHAP attribution, while observation-process features accounted for 10.1% (Figure 2 (4A)). Attribution was distributed across all four clinical domains (Figure 2, 4B). Neurological features accounted for the largest share of total absolute SHAP attribution (34.5%), followed by Respiratory/Acid-Base (22.2%), Metabolic/General (22.0%) and Hemodynamics (21.3%), consistent with the clinical-domain prediction experiments, in which neurological features also produced the strongest isolated-domain performance.

Among the features, the leading cohort-level contributors were GCS motor (12.5% of total absolute attribution), temperature (10.9%), GCS verbal (10.7%), respiratory rate (10.5%), GCS eye (9.0%), and glucose (8.8%) (Figure 2 (4C)). Most attribution for these variables arose from physiological-value features. Glucose showed a more mixed attribution pattern, with 66% arising from physiological-value information and 34% from the observation process.

### 3.7 Patient-Level Contextualized Predictions

Four illustrative held-out episodes were selected to span the four prediction outcome states: one true positive (Case A), one true negative (Case B), one false positive (Case C), and one false negative (Case D) (Figure 3). The cases were used to demonstrate how the framework can reveal how a prediction was formed and what clinical and data context surrounds it, rather than to estimate clinical utility. The calibrated mortality probabilities ranged from 0.1% to 96.0% across the four cases. In all four examples, XGBoost and GRU-D supported the same prediction direction, and the ensemble prediction was far from the operating boundary. Nevertheless, Cases C and D were retrospectively incorrect. This illustrates an important limitation of model context: component agreement and a large decision-boundary margin describe how decisively the models produced a prediction but do not guarantee that the prediction is correct. Case A had a calibrated mortality probability of 96.0%, with hemodynamics contributing the largest domain-level attribution and systolic blood pressure as the leading patient-specific contributor. Recent systolic blood pressure was observed in 19 of 24 hourly intervals and showed a decreasing trajectory, whereas glucose contributed 11.6% of the patient’s XGBoost attribution despite having no measurement during the final 24 hours. Case C had a calibrated probability of 78.9%, but temperature, its largest contributor, had only one observation during the final 24 hours and insufficient observations for estimating a recent trend. Conversely, Case D had a low calibrated probability of 0.9%, with neurological and metabolic variables dominating the explanation, yet the episode was retrospectively a false negative.

**Figure 3.**
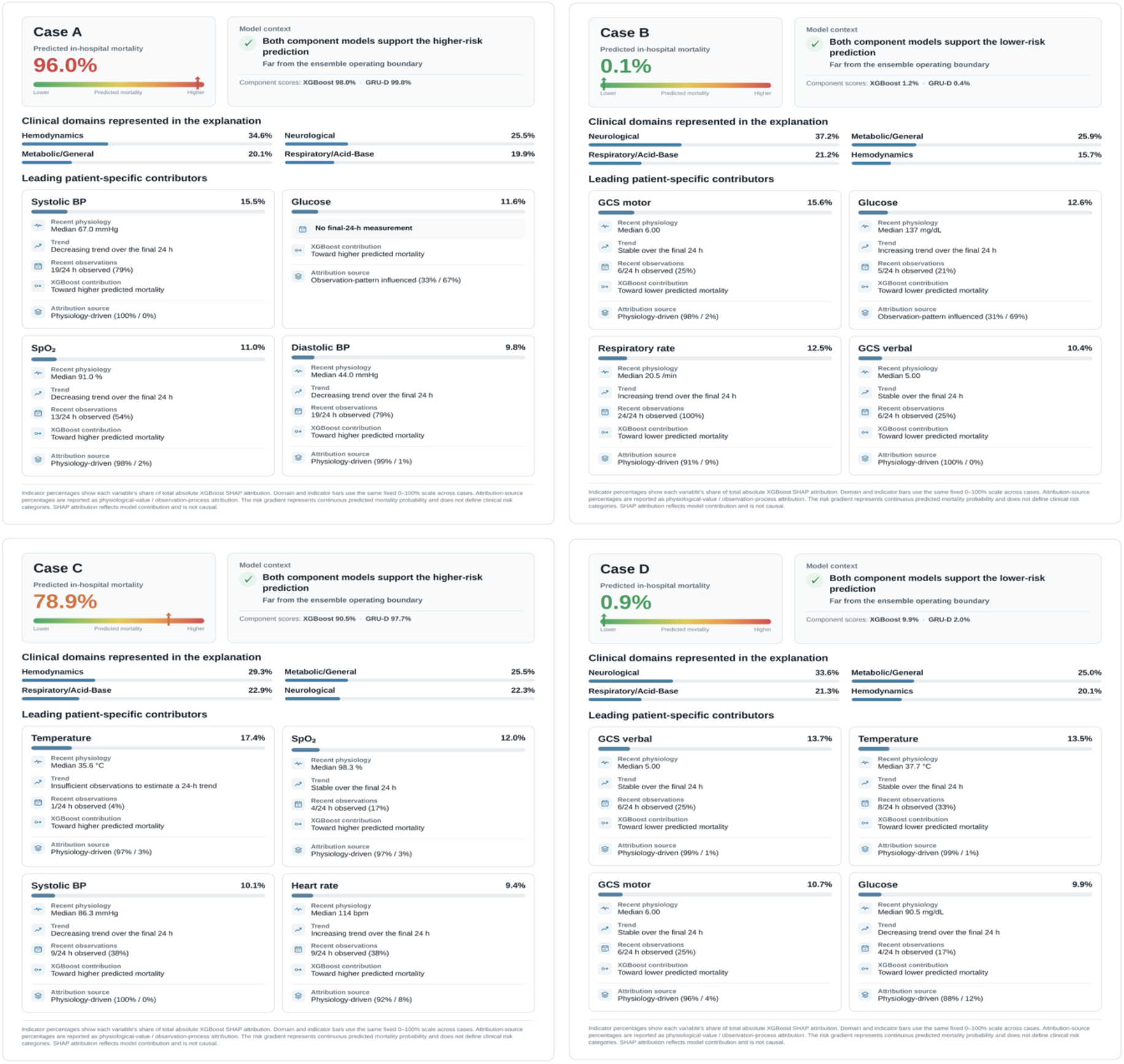
Patient-level contextualized mortality prediction profiles for four representative held-out ICU episodes. Cases A-D represent a true positive, true negative, false positive, and false negative, respectively (outcome correctness is not shown in the cards). Each profile displays the calibrated mortality probability together with component-model context, decision-boundary position, clinical-domain attribution, and the four leading patient-specific XGBoost contributors, including recent physiology, trend, observation frequency, contribution direction, and physiological-value versus observation-process attribution (SHAP values describe model attribution and are not causal).

## 4. Discussion

This study evaluated a multidimensional framework for contextualizing ICU mortality predictions beyond discrimination alone. The weighted ensemble achieved strong held-out performance, but raw probabilities overestimated absolute mortality risk, and validation-based Platt scaling substantially improved calibration without changing discrimination. This reinforces prior work showing that discrimination and calibration capture distinct properties of clinical prediction models^6,7^. Model disagreement and decision-boundary position provided complementary information about how predictions were formed: incorrect predictions showed greater component disagreement and were generally closer to the operating threshold. However, agreement and large margins did not guarantee correctness, as both component models were wrong in a meaningful subset of cases and retrospectively incorrect patient examples could still show concordant, far-boundary predictions. Data and physiological context added further information that was not captured by the risk score. Overall measurement density differed only modestly by correctness, while variable-specific coverage varied substantially across clinical measures. Recent physiological trends were also heterogeneous despite cohort-level medians generally falling within descriptive reference ranges. In the explanation analysis, most XGBoost attribution arose from physiological-value features, but observation-process features contributed 10.1%, consistent with prior work showing that EHR missingness and measurement patterns can encode healthcare-process information in addition to patient physiology^13–15^.

The key contribution is a patient-level contextualization framework that moves beyond presenting a risk score or feature ranking alone. It brings together model agreement and boundary position, recent data availability and physiological trends, and patient-specific attribution around the same calibrated prediction, making the model output more inspectable and clinically contextualized. This direction aligns with clinician-centered work emphasizing that useful explanations should be contextualized within the clinical information surrounding a prediction^11,12^.

This study has several limitations. It was retrospective, used a single-center MIMIC-III benchmark, and did not include external validation or prospective clinician evaluation. SHAP analysis was limited to the XGBoost component, and the disagreement categories, boundary-margin groups, sparse-observation threshold, and descriptive physiological reference ranges were not intended as universal clinical thresholds. The four patient profiles were selected for illustration rather than validation of clinical utility. Therefore, the framework should be viewed as a proof of concept for contextualizing patient-level predictions, not as a validated clinical decision-support intervention. Future work should assess generalizability across institutions and determine, through clinician-centered evaluation, whether these contextual elements improve interpretation, decision quality, and workflow integration without increasing cognitive burden^10–12,16^.

## Data Availability

All data produced are available online at https://physionet.org/content/mimiciv/3.1/

